# HIV-Associated Heart Failure: Phenotypes and Clinical Outcomes in a Safety-Net Setting

**DOI:** 10.1101/2024.05.08.24307095

**Authors:** Matthew S. Durstenfeld, Anjali Thakkar, Diane Jeon, Robert Short, Yifei Ma, Zian H. Tseng, Priscilla Y. Hsue

## Abstract

**Background:** Human immunodeficiency virus (HIV) is associated with increased risk of heart failure (HF) but data regarding phenotypes of heart failure and outcomes after HF diagnosis, especially within the safety-net which is where half of people with HIV in the United States receive care, are less clear.

**Methods:** Using an electronic health record cohort of all individuals with HF within a municipal safety-net system from 2001-2019 linked to the National Death Index Plus, we compared HF phenotypes, all-cause mortality, HF hospitalization, and cause of death for individuals with and without HIV.

**Results:** Among people with HF (n=14,829), 697 individuals had HIV (4.7%). Persons with HIV (PWH) were diagnosed with HF ten years younger on average. A higher proportion of PWH had a reduced ejection fraction at diagnosis (37.9% vs 32.7%). Adjusted for age, sex, and risk factors, coronary artery disease on angiography was similar by HIV status. HIV was associated with 55% higher risk of all-cause mortality (HR 1.55; 95% CI 1.37-1.76; P<0.001) and lower odds of HF hospitalization (OR 0.51; 95% CI 0.39-0.66; P<0.001). Among PWH with HF, cause of death was less often attributed to cardiovascular disease (22.5% vs 54.6% uninfected; P<0.001) and more to substance use (17.9% vs 9.3%; P<0.001), consistent with autopsy findings in a subset (n=81).

**Conclusions:** Among people with HF who receive care within a municipal safety-net system, HIV infection is associated with higher mortality, despite lower odds of HF hospitalization, attributable to non-cardiovascular causes including substance-related and HIV-related mortality.

**Clinical Perspectives:**

- People with HIV develop heart failure 10 years earlier than people without HIV, with a higher proportion with HFrEF at diagnosis.
- HIV is associated with higher mortality among people with heart failure, primarily due to non-cardiovascular causes including HIV/AIDS and substance use.

**Research Perspectives:**

- The reasons for higher mortality in PWH with HF are not yet fully understood; higher levels of myocardial fibrosis may predispose people with substance use and advanced HIV disease to increased risk of mortality.

## Introduction

Human immunodeficiency virus (HIV) infection is associated with increased risk of heart failure (HF), even in the absence of coronary artery disease, with the highest risk among those with higher viral load and lower CD4 counts (1-3). The prevalence of clinical heart failure among persons with HIV (PWH) is estimated to be 6.5%, with higher rates of subclinical LV systolic and diastolic dysfunction (4). Prior studies within tertiary referral centers and the Veterans Affairs system found that PWH with HF have higher risk of HF hospitalization and mortality compared to people with HF without HIV (5-7). In contrast, among people with incident HF within the Kaiser Permanente integrated care system, HIV was associated with all-cause mortality but not HF hospitalizations or Emergency Department visits (8). Over half of PWH in the United States receive benefits through the Ryan White HIV/AIDS Program, the federal safety-net program for PWH. Outcomes reported within tertiary referral centers or integrated care systems may not generalize to the majority of PWH in the United States.

Therefore, we sought to describe heart failure phenotypes, treatment, outcomes after HF diagnosis among PWH compared to people with HF without HIV within a safety-net system with particular attention to mortality, HF hospitalization, and cause of death.

## Methods

### Study Design & Population

This study was an electronic health record-based cohort study of all individuals with HF who received care within the San Francisco Health Network, the public, municipal safety-net health system of San Francisco, California from April 2001-July 2019.

### Exposure

The primary exposure was infection with HIV prior to diagnosis with HF. HF was manually adjudicated by review of a random subset of EHR records and found to be 97% specific for possible HF and 94% specific for definite HF. We initially classified people with HIV using International Classification of Diseases (ICD-9 or ICD-10) codes alone, but by manual adjudication the specificity was only 60%; 40% were false positives without HIV. To increase specificity, we further classified only those with and ICD code and at least one CD4 count or viral load result as HIV+ which increased the manually adjudicated specificity to 99.8% while maintaining 100% sensitivity among adjudicated charts (0 false negatives among 45 adjudicated with HIV ICD codes and neither CD4 nor viral load).

### Outcomes

The primary outcome was all-cause mortality. To ascertain mortality, we matched individuals with the National Death Index and Social Security Death Index using names, date of birth, and social security numbers. For individuals matched and alive, we censored them at the last search date (12/31/2019). For unmatched individuals, we used the last EHR contact date and recorded vital status. Among known deaths, we linked records with the POstmortem Systematic Investigation of Sudden Cardiac Death (POST SCD Study), an ongoing prospective post-mortem study of presumed (WHO-defined) sudden cardiac deaths in San Francisco (9,10).

Our secondary goals were to characterize HF phenotypes by HIV status including echocardiographic findings (left ventricular systolic function, pulmonary artery systolic pressure or pulmonary hypertension, and valvular disease) and angiography results (obstructive coronary artery disease defined as ≥80% stenosis in a major epicardial vessel or ≥50% left main stenosis).

### Covariates

Medical history was obtained from ICD codes available at the time of HF diagnosis with a 1-year look-back period. Ambulatory medications were obtained from hospital admission and discharge medication reconciliation and thus were not available for those who were not hospitalized. Echocardiographic and cardiac catheterization results were obtained through structured text extraction from clinical reports with manual adjudication and correction of discrepancies. The primary underlying cause of death was extracted from death certificate data and classified based on ICD codes.

### Statistical Analysis

We described differences by HIV status using chi-squared tests for categorical variables (and Fisher’s exact test for autopsy data given limited numbers), t-tests for normally distributed continuous variables, and Wilcoxon rank sum tests for non-normally distributed continuous variables. We used Cox proportional hazards models to estimate adjusted hazard ratios for mortality with censoring for loss of follow up. We used directed-acyclic graphs and expert knowledge to decide potential measured confounders (namely, those potentially associated with risk of HIV and with HF outcomes), such that we included age by sex interaction terms, race/ethnicity, and substance use (alcohol, tobacco, cocaine, methamphetamine, opioids) in adjusted models. Although more likely to be mediators than confounders, given differences in past medical history by HIV status we conducted sensitivity analyses including hypertension, diabetes, chronic kidney disease, COPD, liver disease, and cancer. We conducted additional sensitivity analyses considering HIV variables (CD4 count, viral load), Hepatitis C co-infection, and secular trends (pre and post 2011).

### Approval

The University of California San Francisco Institutional Review Board approved this study and granted a waiver of consent. Results are reported in accordance with the Strengthening the Reporting of Observational Studies in Epidemiology (STROBE) guidelines.

## Results

Among those with HF (n=14,829), 906 (6.1%) individuals an ICD code for HIV and 697 (4.7%) had HIV by ICD code and at least one CD4 count and/or viral load measured. Median follow up was 3.5 years after diagnosis with HF (IQR 0.9, 8.3). On average, PWH were 10 years younger at diagnosis. Compared to people without HIV with HF, a higher proportion of PWH were male and had documented substance use (Table 1, p<0.001 for each). Among PWH, the median nadir CD4 count was 133 (IQR 44, 275), and CD4 count at HF diagnosis was 356 (IQR 173, 566).

**Table 1.**
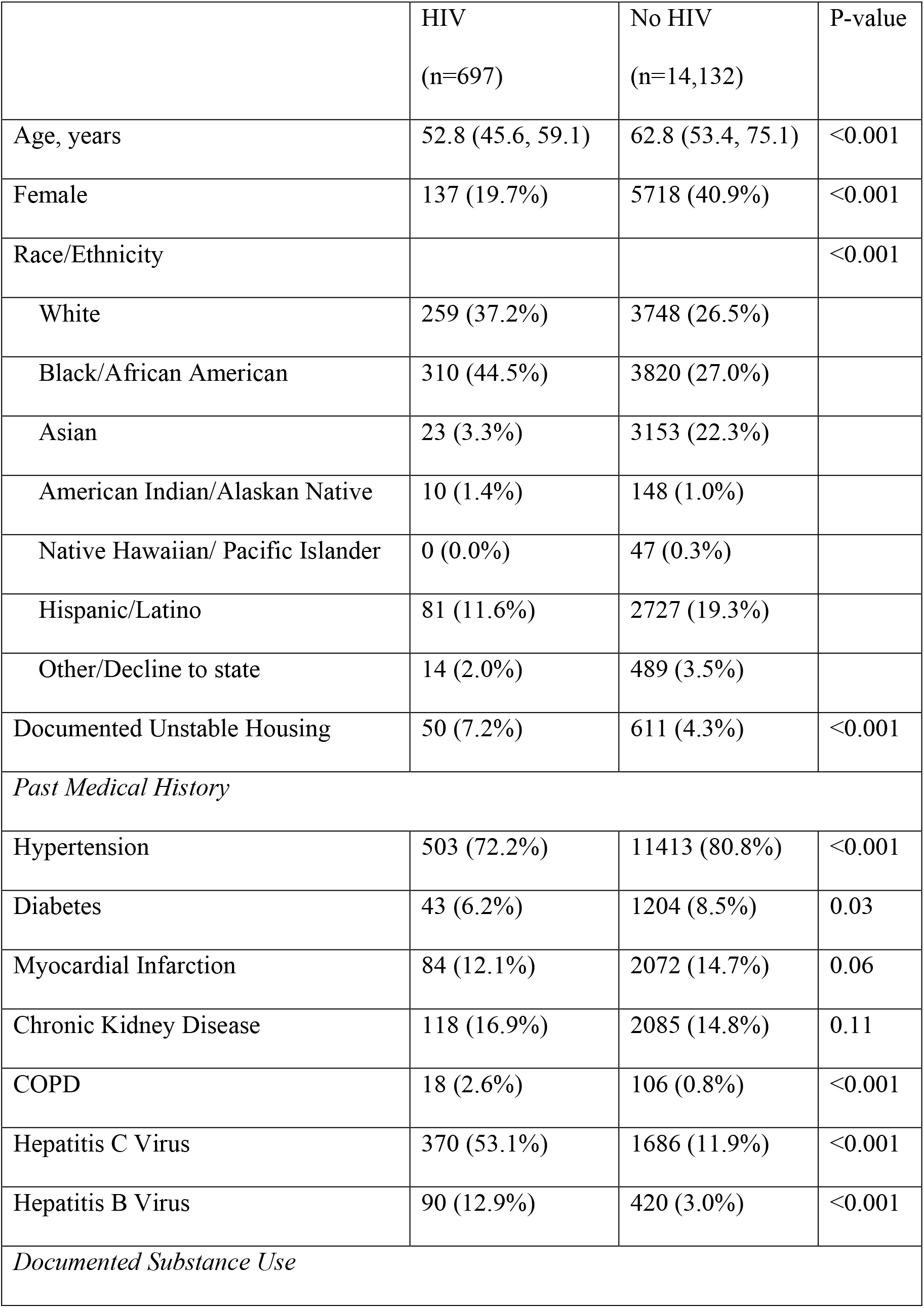
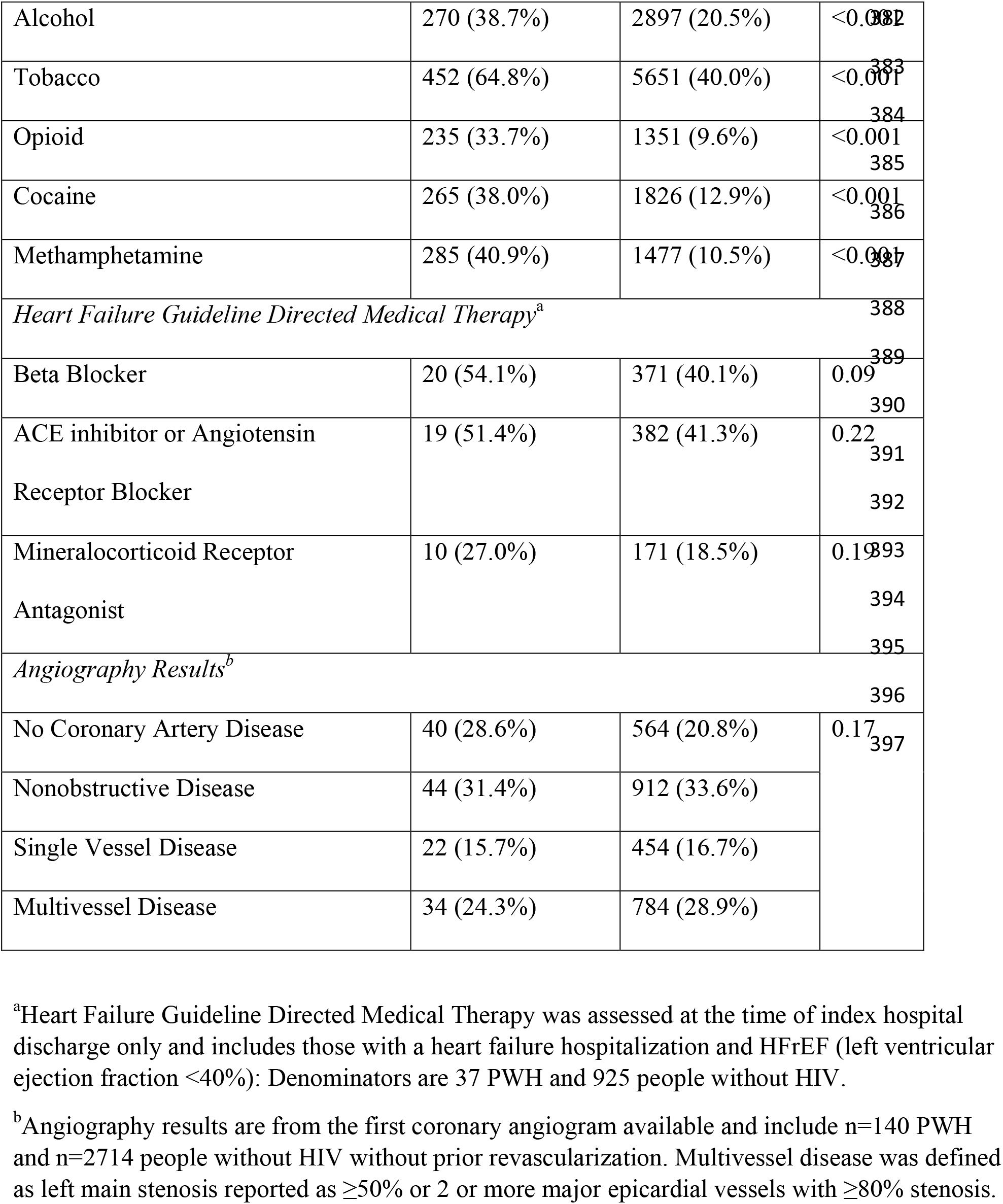
Study Population by HIV Status.

### Echocardiographic Findings

Among those with HF, 593 PWH (85%) and 10,579 without HIV (75%) completed at least one transthoracic echocardiogram linked to their EHR record (Table 2). Using an LVEF cutoff of 40% (or moderately reduced if LVEF was missing) on the first echocardiogram after diagnosis, 225 (37.9%) of PWH had HF with reduced ejection fraction (HFrEF) compared to 2,465 (32.7%) without HIV (p=0.009). The proportion with severely reduced LVEF did not differ significantly by HIV status: 23.0% with HIV versus 20.7% without HIV (p=0.59 for ordinal comparison). Mean LVEF and pulmonary artery systolic pressure at diagnosis were similar between groups (p=0.13 and p=0.77, respectively). A higher proportion with HIV had severe mitral regurgitation (10.5% with HIV vs 6.8% without; p=0.01), but other valvular disease including endocarditis did not vary by HIV status.

**Table 2.** Echocardiographic Findings Among those Who Completed an Echocardiogram.

| Finding | HIV (n=593) | No HIV<br>(n=10579) | P-value |
| --- | --- | --- | --- |
| Echo within 30 days of diagnosis | 313 (52.8%) | 5968 (56.4%) | 0.08 |
| Echo within 6 months of diagnosis | 373 (62.9%) | 7099 (67.1%) | 0.03 |
| HFrEF<sup>a</sup> | 225 (37.9%) | 3459 (32.7%) | 0.008 |
| First EF After HF Diagnosis, % | 45.7 (15.7) | 44.4 (15.6) | 0.13 |
| LV Inner Diameter at End Diastole, cm | 5.1 (0.9) | 5.0 (1.0) | 0.20 |
| *Qualitative LV Function* |  |  |  |
| Hyperdynamic | 11 (2.2%) | 156 (1.7%) | 0.59 |
| Normal | 214 (42.7%) | 4010 (44.2%) |  |
| Borderline | 22 (4.4%) | 439 (4.8%) |  |
| Mildly Reduced | 47 (9.4%) | 842 (9.3%) |  |
| Mild to Moderately Reduced | 17 (3.4%) | 320 (3.5%) |  |
| Moderately Reduced | 50 (10.0%) | 779 (8.6%) |  |
| Moderate to Severely Reduced | 23 (4.6%) | 603 (6.6%) |  |
| Severely Reduced | 115 (23.0%) | 1879 (20.7%) |  |
| Not reported | 2 (0.4%) | 46 (0.5%) |  |
| Regional Wall Motion Abnormalities | 71 (31.1%) | 1023 (34.3%) | 0.33 |
| *Pulmonary Hypertension* |  |  |  |
| Severe Pulmonary Hypertension | 7 (2.2%) | 176 (2.9%) | 0.46 |
| PASP, mm Hg, mean (SD) | 37.1 (16.8) | 36.9 (15.4) | 0.85 |
| *Valve Disease<sup>b</sup>* |  |  |  |
| Severe Mitral Regurgitation | 33 (10.5%) | 406 (6.8%) | 0.01 |
| Severe Tricuspid Regurgitation | 35 (11.2%) | 509 (8.5%) | 0.10 |
| Endocarditis | 5 (0.7%) | 54 (0.4%) | 0.17 |

Notes: a. HFrEF defined as LVEF<40% or for those missing LVEF, qualitative LV function at least moderately reduced. b. Fewer than 1% in each group had severe aortic stenosis, severe aortic regurgitation, and severe mitral stenosis with no differences by group so we did not include them in the table. Abbreviations: HFrEF=Heart Failure with Reduced Ejection Fraction; EF=Ejection Fraction; LV=Left Ventricular

### Ischemic vs Nonischemic Cardiomyopathy

By HIV status, a similar proportion had prior myocardial infarction documented (12.1 PWH vs 14.7% without HIV, p=0.06). Likewise, a similar proportion without prior revascularization completed coronary angiography: 140 (20%) with HIV and 2,714 (19%) without HIV. Angiographic severity of coronary artery disease did not vary by HIV status (p=0.17, Table 1). Adjusted for age, sex, race/ethnicity, comorbidities, and substance use, there remained no difference in angiography results (OR 0.97; 95% CI 0.70-1.34; p=0.87). HIV was not associated with differences in the proportion revascularized (10.2 vs 9.9%; OR 1.12; 95% CI 0.86-1.46; p=0.39) or revascularization strategy (coronary artery bypass graft surgery versus percutaneous coronary intervention; p=0.66).

### HF Admission, Medical Therapy, and Device Therapy among those Hospitalized for Heart Failure

A lower proportion of PWH were hospitalized for HF compared to people without HIV (7.7% versus 11.3%, p=0.001). HIV was associated with lower adjusted odds of HF hospitalization (OR 0.51; 95%CI 0.39-0.66; p<0.001). HIV was not associated with all-cause readmission (OR 0.96; 95%I 0.55-1.67; p=0.89) and there was a trend toward lower odds of HF-specific readmission (OR 0.56; 95%CI 0.31-1.01; p=0.06). Among those with HFrEF who were hospitalized, HIV was associated with higher prescription of HF guideline-directed medical therapy defined as at least one of angiotensin-converting enzyme inhibitor, angiotensin receptor blocker, beta blocker, or mineralocorticoid receptor antagonist (53% vs 39%; OR 2.13 95% CI 1.08-4.21; p=0.03). HIV was associated with similar rates of statin prescription that was not statistically significant after adjustment (35% vs 27%; OR 1.77; 95%CI 0.91-3.44; p=0.09). PWH were also more likely to have outpatient follow up within 30 days of HF discharge (80% vs 65%, p=0.001). Only four PWH had defibrillators implanted, two each for primary and secondary prevention: one each were for cardiac resynchronization therapy.

### Mortality

In total, 41% died before the end of follow-up time including 354 (50.8%) with HIV and 5,784 (40.9%) without HIV (p<0.001). HIV was associated with higher age-, sex-, and substance use-adjusted risk of all-cause mortality (HR 1.55; 95%CI 1.37-1.76; p<0.0001; **Central Illustration**). The median survival after HF diagnosis was 6.1 years (95% CI 5.4-7.1 years) among PWH compared to a median survival of 11.3 years (95% CI 10.7-11.7) years among people without HIV. The higher risk remained elevated among PWH accounting for past medical history (HR 1.37; 95% CI 1.22-1.54; p<0.001). Results were similar for 1-year mortality with adjusted relative risk ratios of 1.72 (95% CI 1.50-1.97) adjusted for age, sex, and substance use, and 1.47 (95% CI 1.28-1.70) additionally accounting for past medical history. The absolute risk difference in 1-year mortality among those with HIV compared to without HIV was 11.0% (95% CI 7.5%-14.4%) and 7.3% (95% CI 4.1-10.4) accounting for other past medical history.

Both lower nadir CD4 and lower CD4 at the time of HF diagnosis were associated with worse survival among PWH: HR 1.37 for nadir CD4 count < 200 (95% CI 1.08-1.75; p=0.009) and HR 1.69 for current CD4 count<200 (95% CI 1.34-2.13; p<0.001), respectively. Viral load less than 500 copies/ml was not associated with mortality (HR 1.02; 95%CI 0.80-1.31; p=0.87). There was not a significant interaction with Hepatitis C coinfection (p_interaction_=0.53). Restricting the PWH to only those virally suppressed (<75 copies per milliliter, n=76) compared to people without HIV yielded similar adjusted hazard for mortality (HR 1.52, 95% CI 1.27-1.83; p<0.001).

In an exploratory analysis to consider secular trends, we divided the study into 2001-2010 and 2011-2019. Among those diagnosed with HF more recently, HIV was associated with a smaller increase in mortality relative to people without HIV (HR 1.24, 95% CI 1.02-1.51; p=0.03) compared to before 2011 (HR 1.75, 95% CI 1.52-2.01, p<0.001, p_interaction_=0.004).

By death certificate data (n=3650, 262 PWH), there were notable differences by HIV status (**Figure 1**). Among PWH, 36% had the primary underlying cause of death classified as HIV/AIDS-related, 19% as cardiovascular, 16% as due to accident/injury (including overdose). Among those without HIV, 46% of deaths were attributed to cardiovascular causes, 11% to neoplasms and blood disorders, and only 8% to accidents, injuries, and overdoses. PWH had twice the odds of cause of death attribution to overdose or substance use (OR 2.1; 95% CI 1.53-3.00; p<0.001), but not after accounting for age, sex, and documented history of substance use (OR 0.96; 95% CI 0.67-1.37; p=0.82). In contrast, HIV was associated with lower odds of having cardiovascular disease as the primary recorded cause of death (even though all had HF) with an adjusted OR of 0.32 (95% CI 0.23-0.43; p<0.001) accounting for age, sex, substance use, and comorbidities.

**Figure 1.**
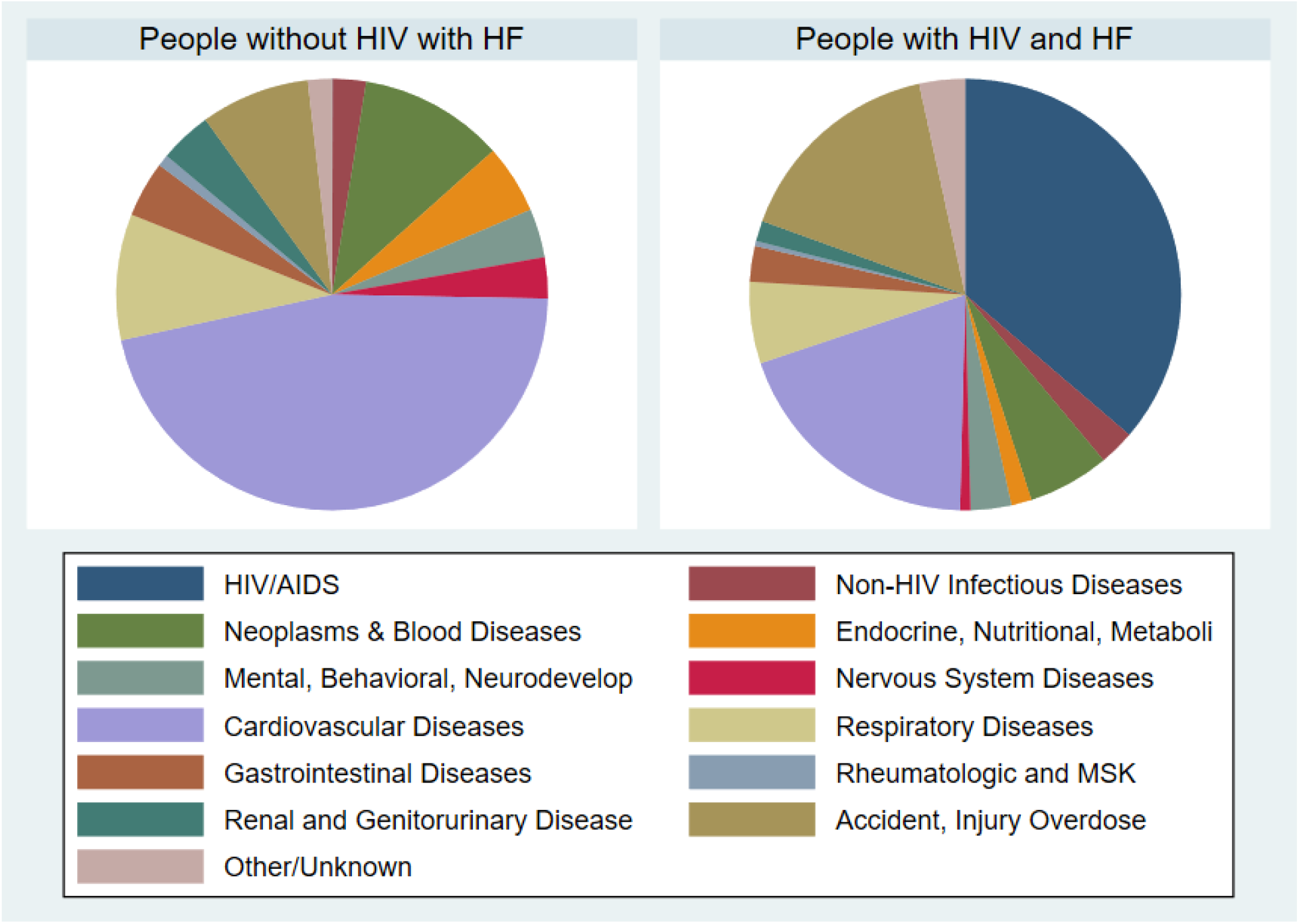
Primary Underlying Cause of Death among People with Heart Failure as Recorded on Death Certificates by HIV Status. Figure Legend: Primary underlying cause of death as reported on death certificates classified by International Classification of Diseases (ICD) codes by HIV status. Liver disease is classified under gastrointestinal diseases.

Autopsy data were available for 81 individuals (14 PWH), who died between 2011 and 2019 in San Francisco County, 51 of whom were adjudicated as part of the POST SCD Study. Of PWH, 4 (29%) met the WHO definition for presumed sudden cardiac death, which excludes those with obvious external cause of death such as trauma or drug paraphernalia at the scene (11), compared to 38 (56.8%) of those without HIV (p=0.08). Among these presumed SCDs, PWH had a trend towards fewer adjudicated arrhythmic causes (21.4% vs 41.8%, p=0.23), and more non-cardiac causes (78.6% vs 50.8%; p=0.08), including occult overdose with toxicology evidence without evidence of drug use at the scene (64.3% vs 37.8% of noncardiac deaths; p=0.12).

## Discussion

In this 18-year study of people with HF within a municipal safety-net system, PWH had a greater all-cause mortality despite lower odds of hospitalization for HF, more often due to non-cardiovascular causes, especially overdose. PWH developed HF a mean 10 years younger than people without HIV, with higher rates of documented substance use. HFrEF was more commonly diagnosed in PWH, but echocardiographic and angiographic findings were similar compared to cases without HIV. Although the mortality gap has narrowed over time, PWH with HF remain at elevated risk compared to people with HF without HIV. These findings have implications for targeted strategies to reduce higher mortality in HF for PWH.

### Similar Associations with Mortality as Compared to Prior Studies

Several prior studies have examined the association of HIV with HF outcomes in very different patient populations. A study of U.S. Veterans from 1999-2018 that included 5,747 PWH and 33,497 controls found higher mortality and hospitalizations among PWH, but the main limitation is that they included 98% men (2,7). They also found that PWH develop HFrEF at younger ages (2). Because the VA study focused on men, another study included ambulatory women within the Partners HealthCare System (now Mass General Brigham) with a primary outcome of incident HF hospitalization (5). Another study included patients hospitalized with acute decompensated HF at Bronx-Lebanon Hospital Center in New York in 2011 with primary outcomes of 30-day readmissions and mortality (6). These studies within tertiary referral centers both found that HIV is associated with HF hospitalization, in contrast to our findings. Our findings are most consistent with a study within the Kaiser integrated care system, where HIV was associated with mortality but not HF hospitalizations or Emergency Department visits, (8). Our study extends these findings to the safety-net where many PWH in the United States receive care.

Our study sheds light on a possible explanation for higher mortality among PWH with HF. From 2000-2009, presumed sudden cardiac death (SCD) accounted for 86% of cardiac deaths among PWH in San Francisco, and antecedent HF was strongly associated with SCD (12). Among these individuals, lower LVEF and diastolic dysfunction were both associated with increased risk of SCD (13). In California from 2005-2015, PWH had 2.5-fold higher risk of out of hospital cardiac arrest (14). One group estimated that the risk of SCD was 10% per year among PWH without indications for primary prevention implantable cardioverter-defibrillators (15). We found very low use of defibrillators, much lower than would be expected for the proportion with severely reduced LVEF. A population-based autopsy study in San Francisco found that PWH were at 2.25 times higher risk of WHO-defined SCD, mostly attributable to higher rates of occult overdose, with 1.87 times higher incidence of arrhythmic death and interstitial myocardial fibrosis on histology (16). Our finding that the primary cause of death is most often attributed to HIV/AIDS suggests that many deaths are non-cardiac even among those with HF. Our findings that HIV is associated with higher risk of substance-related death are confirmed in the autopsy subset and are likely attributable to higher rates of substance use among PWH (17) as the association was attenuated accounting for documented substance use. Rising rates of substance-associated cardiovascular death across the United States suggest that this growing problem extends to the general population without HIV (18).

One potential explanation for lower rates of HF hospitalization specific to the study population may be that PWH may be better connected to ambulatory care within the municipal health system in San Francisco compared to people without HIV, as demonstrated by the higher rates of ambulatory follow-up after HF hospitalization. This may be in part due to the established HIV Cardiology Clinic that is embedded within the main HIV primary care clinic at San Francisco General Hospital. Secondly, among those hospitalized with HFrEF, PWH had higher prescription rates for guideline directed medical therapy compared to people without HIV. Although not conclusive, our study suggests that access to cardiology care does not fully address the increased risk among PWH.

How can we mitigate the increased risk of mortality among PWH with HF? Prevention of myocardial infarction with statins may decrease atherosclerosis and ultimately reduce incident HF among PWH. Earlier detection of Stage B HF (asymptomatic structural heart disease) among PWH could allow for initiation of disease-modifying medical therapy. Biomarkers could identify those at elevated risk (19,20). The MIRACLE HIV trial randomized 40 PWH without known heart disease (Stage A: asymptomatic with risk factors) 1:1 to eplerenone and placebo for 12 months and found that it prevented worsening of myocardial perfusion and function (21). Animal models suggest that immune therapy targeting inflammation may prevent cardiomyopathy (22), although this has not been tested in humans. Most important, however, may be providing high-quality HIV and primary care to PWH even among those with HF, with particular attention to substance use treatment and overdose prevention.

### Limitations

The major limitation of this observational EHR-based study is the risk of residual confounding which limits causal interpretation—PWH may differ from those without HIV in ways that affect HF outcomes not captured in billing codes. Some covariates are notably imperfect such as unstable housing and substance use; other socioeconomic variables including income, education, primary language, immigration status, and current housing were not measured. Secondly, we did not have time-updated covariates, so we used baseline covariates assessed at the time of HF diagnosis. Ambulatory medication data were only available for those who were hospitalized through medication reconciliation, so we were unable to assess use of antiretroviral therapy or titration of GDMT. Previous studies have suggested that ICD coding has a low specificity for HF (23), but in our system manual adjudication demonstrated a high specificity. Emergency department and urgent care visits and specialty provider type for inpatient physicians were not available, limiting our ability to assess acute care utilization. Finally, recorded cause of death is a poor measure of actual cause of death, but we confirmed our findings by linking records with the postmortem data available in the POST SCD Study.

## Conclusions

Among people with HF who receive care within a municipal safety-net system, HIV is associated with higher all-cause mortality but not HF hospitalization. In the current era of antiretroviral therapy, HF phenotypes between those with and without HIV are similar, although PWH develop heart failure a decade younger and are more likely to present with HFrEF. As recorded on death certificates and found on autopsy, non-cardiovascular causes of death including substance-related, and HIV/AIDS deaths are predominant among PWH. Further research is needed to identify and test strategies to mitigate the increased risk of mortality among PWH with HF including addressing high rates of substance use and treatment of HIV.

## Data Availability

Deidentified data is available upon reasonable request to the corresponding author for non-commercial, scientific purposes.

## Abbreviations

HF: Heart Failure
HFrEF: Heart Failure with Reduced Ejection Fraction
HIV: Human Immunodeficiency Virus
PWH: persons with HIV

---

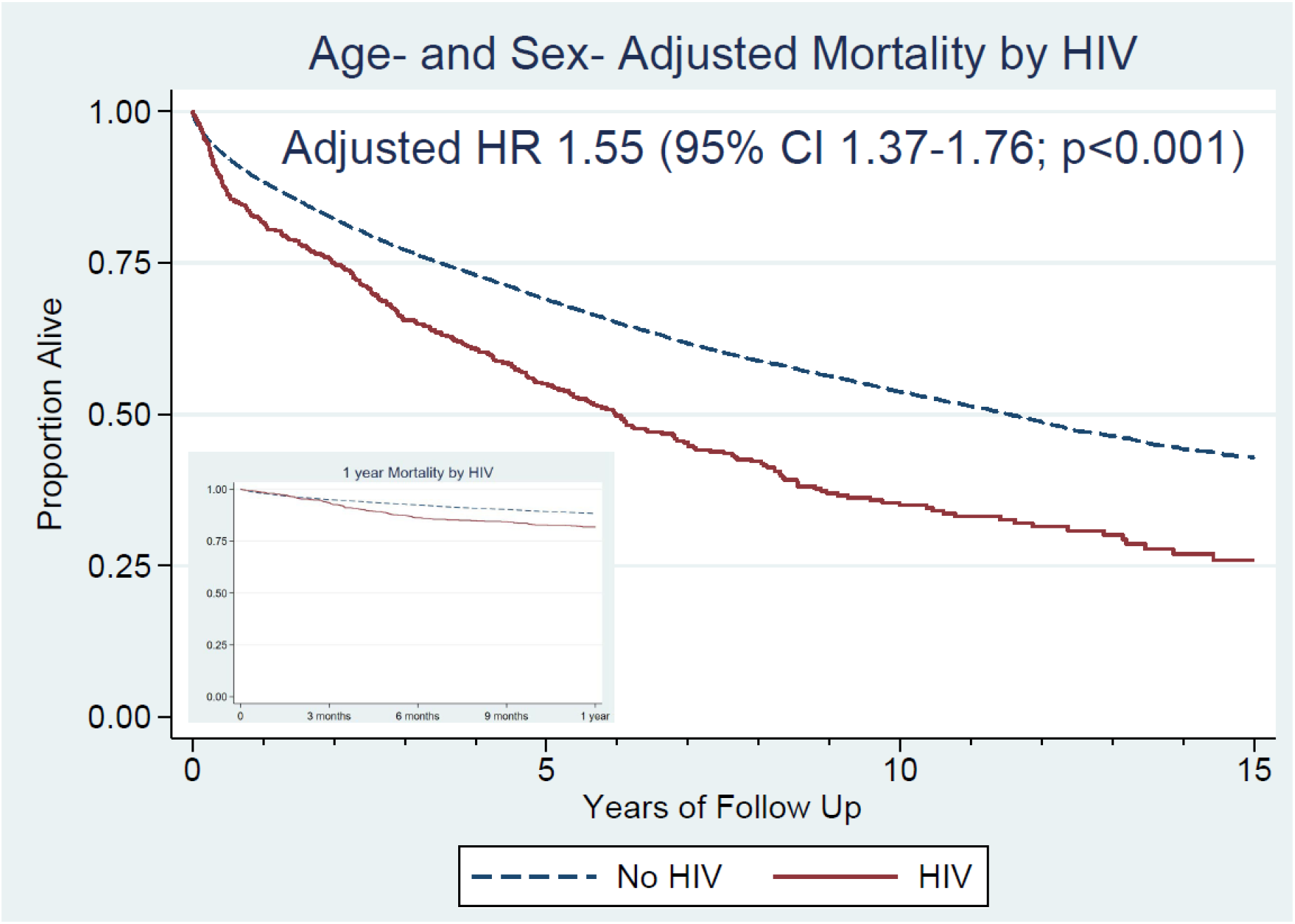
Central Illustration. Kaplan-Meier and Adjusted Survival Plots by HIV. Age and sex adjusted survival curves for people with and without HIV from index time of heart failure diagnosis up to 15 years of follow up (main figure) and from 0-12 months (inset). Survival curves separate about 3 months after HF diagnosis (inset), with an adjusted hazard ratio of 1.55 (95%CI 1.37-1.76; p<0.0001).

